# Menstrual Hygiene Management Practices among Girls and Women after the Chhaugoth Demolition Campaign in Chaurpati Rural Municipality, Achham, Nepal

**DOI:** 10.1101/2025.10.28.25339015

**Authors:** Kalpana Jnawali, Prativa Tiwari, Ayusha Ghale, Jagat Prasad Upadhyay, Anjana Sigdel, Sapana Thapa, Anushka Shrestha, K.C. Kiran, Sirjana Pandit Pahari, Damaru Prasad Paneru

## Abstract

**Background:** Menstruation, also called Chhaupadi (in Sudurpaschim and Karnali provinces); is extricable connected to social taboos and stigma in Nepal. During menstrual periods and after child birth, girls and women are traditionally bound to stay for certain days in huts called Chhaugoth. Such practices have adverse social, economic and health consequences. It is legally punishable in Nepal; however, such practices remain prevalent. Recently, the Government of Nepal launched the Chhaugoth demolition campaign to eliminate such practices. In this context, this study aimed to identify the menstrual hygiene management practices after Chhaugoth Demolition Campaign in Chaurpati Rural Municipality Achham district of Nepal.

**Methods:** This was a cross-sectional quantitative study; conducted among 385 resident girls and women of the Chaurpati Rural Municipality who were in the menstrual life span (menarche to pre-menopause). A multistage sampling technique was used to select respondents from the different wards of Chaurpati Rural Municipality. Data were collected through individual interview using KOBO toolbox. Data were analyzed using IBM’s SPSS 21. Ethical approval was obtained from IRC of Pokhara University. Appropriate descriptive statistics such as mean/median and SD were applied.

**Results:** Most women and girls (93.5%) reported that the existing Chhaugoths were demolished. Likewise, majority (85.5%) now reside in separate rooms at home during menstruation after Chhaugoth demolition campaign; however, about 11% still use Chhaugoths, often covertly. Girls and women reported improved living arrangements (30.8%); however, menstrual hygiene management practices remained considerably poor. While, 48.6% used sanitary pads but the practice of waste disposal remained suboptimal, with 29.1% burning and 15.1% dumping directly in water resources posing environmental health risks.

**Conclusion:** This study demonstrates progress in shifting women from hazardous menstrual seclusion to safer, in-house accommodations during menstruation. However, critical gaps persist in menstrual waste disposal and supportive infrastructure. To achieve sustainable menstrual dignity and equity, interventions must combine targeted behavior change communication with early, pre-menarche education, fostering a generational shift in practices and norms.

## INTRODUCTION

Menstruation is the periodic physiological process; often stigmatized and linked to social taboos, and harmful practices around the world, especially in low and middle income countries of South Asia (1,2). Although, menstruation being a natural phenomenon, cultural taboos associated with misconception often portrays menstruating women as impure, or unclean, which leads them to face widespread exclusion in social, educational and religious activities in many South Asian countries (3,4). In Nepal, many menstruating women experience and follow the practice of Chhaupadi, a form of menstrual exile in secluded huts known as Chhaugoths (5,6). Chhaupadi is derived from two words: ‘Chau’ meaning ‘menstruation’ and ‘padi’ meaning ‘women’ (7) where women are forced to isolate and sleep inside a small shed/hut known as Chhaugoth (5). Despite the legal prohibition under the 2017 criminal code of the Government of Nepal, this ill rooted tradition runs deep within the Nepalese society, particularly in Karnali and Sudurpaschim Provinces; including in Achham districts of Nepal (8,9).

Achham district has one of the highest number of Chhaugoths, which is deeply influenced by the rooted Hindu belief that menstruation is impure (1,10). These types of cultural norms and practices dictate that women and girls must isolate themselves during their menstruation cycle, and as failing to do so will anger deities and bring misfortune upon their families and communities (1). These ill practices have devastating consequences, where numerous reports have documented the deaths of women and girls due to snakebites, suffocation, sexual violence, fire, hypothermia while confined in Chhaugoth (8,11,12). Tragically, recent incidents of death of women and girls residing in the Chhaugoth while adhering to the deadly practice (13,14) underscoring the urgent need for culturally sensitive public health interventions.

Globally, Menstruation hygiene management (MHM) is shaped and experienced in diverse ways by individuals, influenced by different factors such as personal choice, access to resources, economic and social status, cultural and religious beliefs, traditional taboos, education levels, and personal attitudes towards menstruation (15,16). Over 500 million women and girls lack access to adequate menstrual hygiene materials, and research shows that many still face shame, embarrassment, and stigma during menstruation (17–19). In many low and middle-income countries (LMICs), limited access to toilets, clean water, safe menstrual products, and menstrual health education, many girls miss school or leave altogether (20,21).

Unsafe menstrual hygiene practices often shaped by socio-cultural taboos, poverty, and limited access to education and health resources. Moreover, in remote or unsafe environments like Chhaugoths, Achhami women and girls also face increased risks of snake or insect bites, wild animal attacks, sexual violence, and even death from suffocation or exposure (8,11,12,22). Recognizing menstrual health and hygiene as a part of the fundamental right to health and human dignity, the Government and international organizations actively promoting safe practices; yet, stigma, silence, and lack of access to menstrual hygiene resources continue to undermine gender equity and the right to reproductive health (23,24). United Nations Children’s Fund (UNICEF), World Health Organization (WHO), and numerous non-governmental organizations (NGOs) have introduced and implemented multitude of interventions focusing on education, menstrual safety products distribution and water, sanitation, and hygiene (WASH) infrastructure building to support the dignity and health rights of women and girls (25).

In May 2005, the Supreme Court of Nepal recognized Chhaupadi as discriminatory and dangerous custom (5). However, for many years Chhaupadi was decriminalized because of the absence of specific legislation until decades later change only came in 2017. Then the Government of Nepal issued an eight-point circular to the local authorities and administration in 19 districts across Sudurpaschim and Karnali provinces with the active mobilization of local leaders, civil society and media for demolition of Chhaugoths (26–28). Despite these bold interventions, the real impact on the ground remains unclear, as there was no formal monitoring mechanism to assess the post campaigns outcomes. There still lacks evidence regarding whether these efforts led to meaningful improvements in menstrual hygiene practices at community level. Consequently, post campaign status of Chhaupadi did not revealed publicly. Furthermore, limited studies have examined the lived experiences of women in aftermath of anti-Chhaugoths initiatives; nonetheless, the realities whether legal reforms have created more dignified and safer menstrual health practice or not.

Although, healthy menstruation is increasingly recognized as a key indicator of overall health, dignity, and reproductive well-being, it remains entangled with harmful traditional practices such as Chhaupadi, a form of menstrual seclusion that requires girls and women to stay in Chhaugoths during their menstrual period in many rural areas of western Nepal. This study aims to assess the current living arrangements of menstruating girls and women and examine menstrual hygiene management (MHM) practices in Chaurpati Rural Municipality of Achham.

## Materials and Methods

This was a cross-sectional quantitative study; conducted to assess the menstrual hygiene management (MHM) practices of girls and women (participants) after Chhaugoth demolition campaign (2017 and subsequent events). The study was conducted in Chaurpati Rural Municipality of Achham district, Nepal. This Municipality is administratively divided into 7 Wards and multiple sub-units called villages/tole. This area was selected due to reported ongoing prevalence of the Chhaupadi practice and its inclusion in the government’s demolition campaigns. Girls and Women who were in the menstrual life span (menarche to pre menopause) were the participants of study. The sample size of 385 participants was determined using Solvin’s formula with a 5% margin of error, assuming a prevalence of Chhaupadi practice (p=50%) with the 95% confidence level. Data collection and participant recruitment took place from 26 May 2024 to 9 June 2024. A multistage sampling technique was employed: first stage-alphabetical listing of all villages was prepared and then, 50% of these villages were randomly selected. Ward number 7 was excluded due to the lack of evidence of the demolition campaign. In the second stage, number of participants to be included per ward were estimated proportionately and then participants were selected from each of the selected villages using quota sampling. Eligible participants meeting the eligibility who provided the consent to participate were included in the study until the desired samples achieved. Face to face interview was conducted with the participants to gather information on socio-demographic characteristics, living arrangements and MHM practices. Similarly, an observation checklist was used to assess the physical conditions of living place used during menstruation and/or to supplement the information. Data was collected using KOBO toolbox. Tools were developed in both English and Nepali Language. Trained enumerators collected data. A pretesting of the tool done among 10% sample participants of Sudurpaschim Province who were currently living in Pokhara metropolitan and/or studying in different institutes of Pokhara.

Collected data were exported to IBM SPSS 21 for analysis. Descriptive statistics (frequency, percentage, mean, median, minimum, maximum, standard deviation) were computed to describe the participant’s sociodemographic characteristics and MHM practices. Current menstrual hygiene management practices were assessed of key practices such as the type of menstrual products, frequency of changing menstrual materials, cleaning and disposal practices, and personal hygiene behaviors.

Ethical approval was obtained from the Institutional Review Committee (IRC) of Pokhara University (Ref no: 151/2080/2081.); and approval was also obtained from the office of Chaurpati Rural Municipality. Written consent was obtained from all the participants prior to data collection. Assent was obtained for participants under 18 years old from their parents/guardians. Confidentiality and anonymity of the participants was maintained throughout the process.

## Results

A total of 385 women/girls from six different wards were included this study, where 21.8% participants were from Ward no.1; followed by others (table 1). Nearly half of the women/girls (42.9%) were aged 20-29 years (Mean age: 28.75±8.49 years). Majority of them were Chhetri/Brahmin (70.7%), with nearly all of the women/girls were Hindus (98.7%). Four out of every five (80.5%) participants were married (80.5%); with more than half lived in Joint family (54.5%). Nearly two third participants were Housewife (64.4%) and the average monthly income was 107.17 USD (Oct 3,2025). Additionally, more than a quarter of the participants had Secondary education (29.9%) and Informal Education (28.6%) and few had Bachelor’s, Maters and higher degree. (**Table 1**)

**Table 1:** Sociodemographic characteristics of the respondents.

| Sociodemographic characteristics | Frequency (n=385) | Percentage |
| --- | --- | --- |
| **Residence (Ward-wise)** |  |  |
| 1 | 84 | 21.8 |
| 2 | 77 | 20.0 |
| 3 | 56 | 14.5 |
| 4 | 66 | 17.1 |
| 5 | 55 | 14.3 |
| 6 | 47 | 12.2 |
| **Age (in completed years)** |  |  |
| Less than 20 | 59 | 15.3 |
| 20-29 | 165 | 42.9 |
| 30-39 | 104 | 27.0 |
| 40 or above | 57 | 14.8 |
| (Mean age: 28.75±8.49 years; Min-Max: 13-50 years) |  |  |
| **Caste/ Ethnicity** |  |  |
| Brahmin/ Chhetri | 272 | 70.7 |
| Dalit | 103 | 26.8 |
| Janjati | 1 | 0.3 |
| Others (unclassified) | 9 | 2.3 |
| **Religion** |  |  |
| Hindu | 380 | 98.7 |
| Christian | 5 | 1.3 |
| **Marital Status** |  |  |
| Married | 310 | 80.5 |
| Unmarried | 63 | 16.4 |
| Widow | 12 | 3.1 |
| **Family type** |  |  |
| Joint Family | 210 | 54.5 |
| Nuclear Family | 175 | 45.5 |
| **Occupation** |  |  |
| Housewife | 248 | 64.4 |
| Student | 58 | 15.1 |
| Farmer | 47 | 12.2 |
| Others (Daily wage worker, Laborer, Service holders) | 24 | 6.2 |
| Business | 8 | 2.1 |
| **Monthly Household Income (in USD) : Mean: 107.17 USD; Median: 70.42 USD; Range (Min-Max): 7.04-1056.56 ( 1 USD=142 NRs; exchange rate on date: Oct 3,2025)** |  |  |
| **Education level** |  |  |
| Bachelor's level and above | 16 | 4.2 |
| Secondary level | 115 | 29.9 |
| Basic level | 86 | 22.3 |
| Informal Education | 110 | 28.6 |
| Illiterate | 58 | 15.1 |

*#Brahmin/Chhetri refers to respondents from higher Hindu groups traditionally occupying priestly and warrior classes. Dalit includes respondents from marginalized groups. Janjati represents indigenous ethnic groups, while Others includes participants not classified under the above categories.*

Over the three quarters of the participants (75.8%) had the experience of staying in a Chhaugoth during menstruation. Before Chhaugoth elimination, nearly three quarters of the participants (74.8%) stayed in Chhaugoth. More than a third of the participants stayed in Chhaugoth for 4 days (35.1%), followed by 5 days (27.8%). More than a third of the participants used warm clothes (35.8%), followed by Local matrices (Moto Gadda) (25.5%), and few participants used straw materials or bare floor as a sleeping materials. After the Chhaugoth demolition campaign, a great majority (85.5%) of the participants now stay in the separate room at home; however, 11.4% of the participants were staying in Chhaugoths. Among those who still stay in Chhaugoths during menstruation, nearly three quarters (72.7%) stayed for 4 days. Three-fifth participants (60.5%) now use warm clothes, followed by thick local mattresses (36.6%) and 0.3% used bare floor for sleeping during menstruation. Majority of the participants (78.2%) still faces restrictions followed during menstruation even after the Chhaugoth demolition campaign; indicating that the Chhaupadi tradition is still prevalent in the society. Almost all the participants (93.5%) reported that their Chhaugoths were demolished by the government’s campaign. (**Table 2**)

**Table 2:**
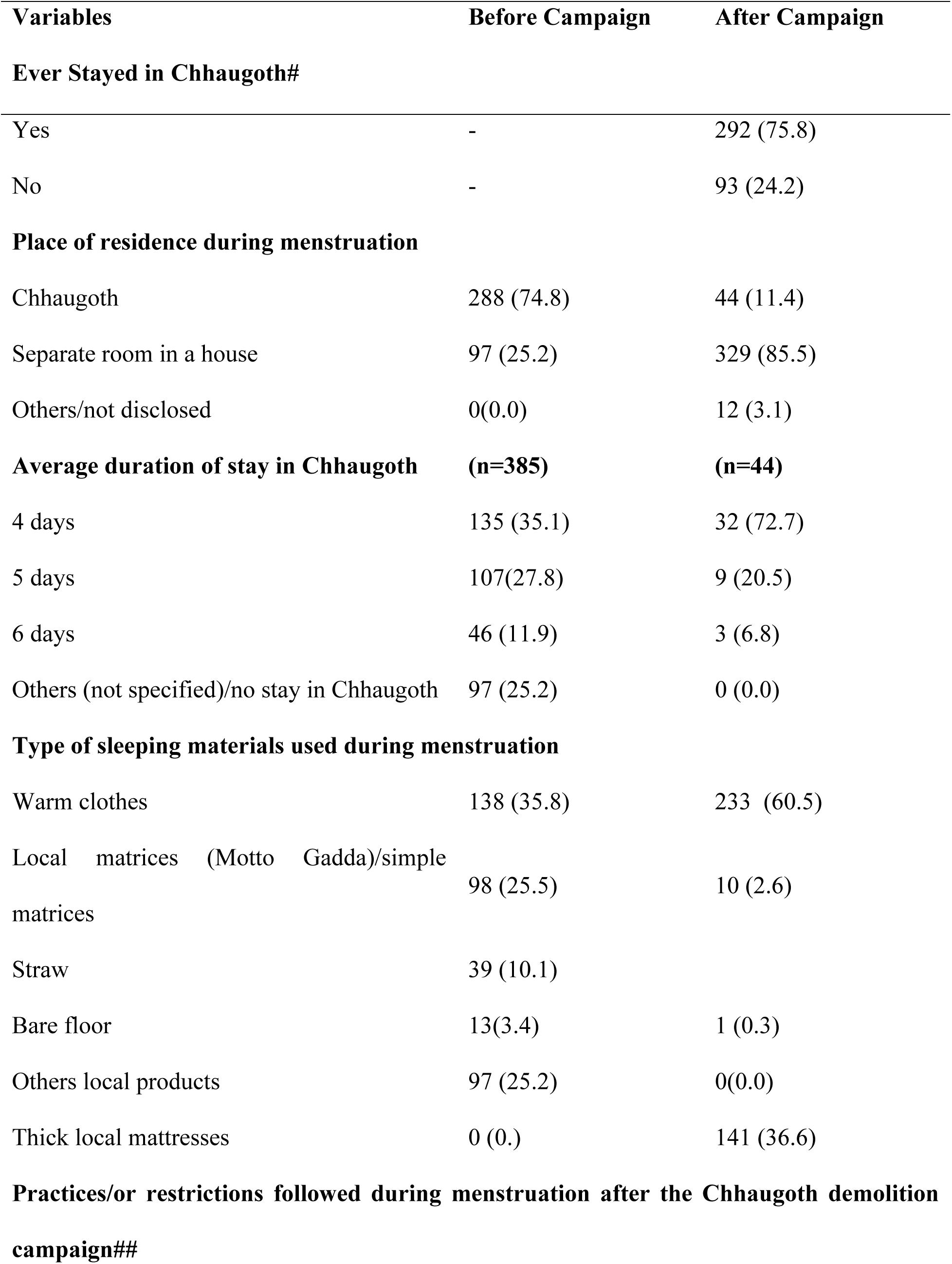
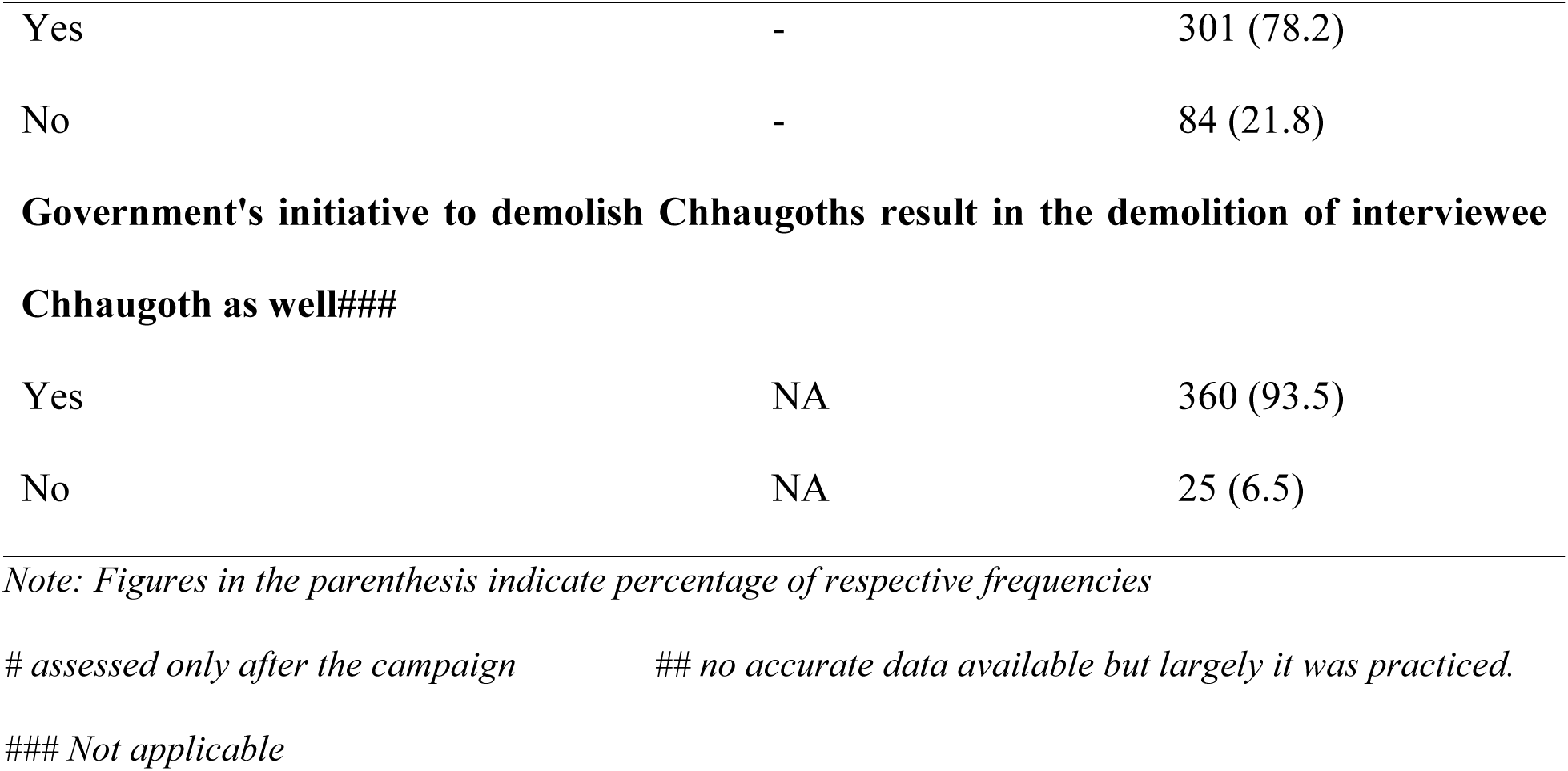
Menstrual Living Arrangements - Practices before and after Chhaugoth Demolition Campaign *(n=385)*

Nearly half of the participants (48.6%) used Sanitary pads. Likewise, more than one-third (37.1%) of the participants used old cloth rag, followed by a small minority of the respondents used locally cloth pads, menstrual cups as menstrual hygiene products. Nearly two thirds of the respondents (61.6%) participants reported that these menstrual hygiene products were easily accessible. Additionally, over half of the participants (52.2%) had received education or training on safe Menstrual Hygiene Practices. Similarly, half of the respondents (50.6%) observed changes menstrual hygiene practices after the Chhaugoth demolition campaign. The observed changes were improved living conditions, improvements in their hygiene and sanitary conditions, and reduced stigma followed by use and disposal of pads and access to healthy diet (table 3). Likewise, majority of the participants (87.8%) reported that they no longer stay in Chhaugoth after the elimination campaign. Almost one-third of the participants (32.5%) changed their sanitary products as per need, followed by more than a quarter (26%) changed twice a day. Majority of the respondents (83.6%) reported that they always have access to clean water and soap, while only few lacked access to both clean water and soap during menstruation. Nearly a third of the respondents (29.1%) burned their menstrual waste, followed by a quarter of respondents (25.6%) reused after washing, while other minority either dumped into river stream or in pit or an open area to burn it. Almost two thirds of the respondents (61.8%) felt comfortable sharing and discussing menstruation issues. Likewise, when asked about whom they share such issues Nearly half of the respondents (46.2%) shared their menstrual related problems with their friends, followed by more than a third of the respondents (35.7%) to their family members and 18.1% to the Health workers. Furthermore, almost all of the respondents (99.7%) bathes during menstruation, where more than half and quarter of the respondents bathed either in their first and second day. Additionally, nearly half of the respondents take their bath for one day (43.6%) and regularly for three days (41.6%) while only a small minority (14.8%) bathed regularly for two days after their first bath during menstruation. Nearly three quarters of respondents (71.7%) responded using shared toilet, while other proportion of the respondents either used open space, or separate toilet for defecation and urination during their menstruation. Regarding cleaning of their private parts, almost all of the respondents (99.5%) clean their private parts during menstruation. Also, more than one-third of the respondents (38.4%) reported cleaning genitalia more than two times daily during menstruation, where more than half of the respondents (53%) used soap and water, followed by (42.1%) used water only to clean their genital area during menstruation. (**Table 3**)

**Table 3:**
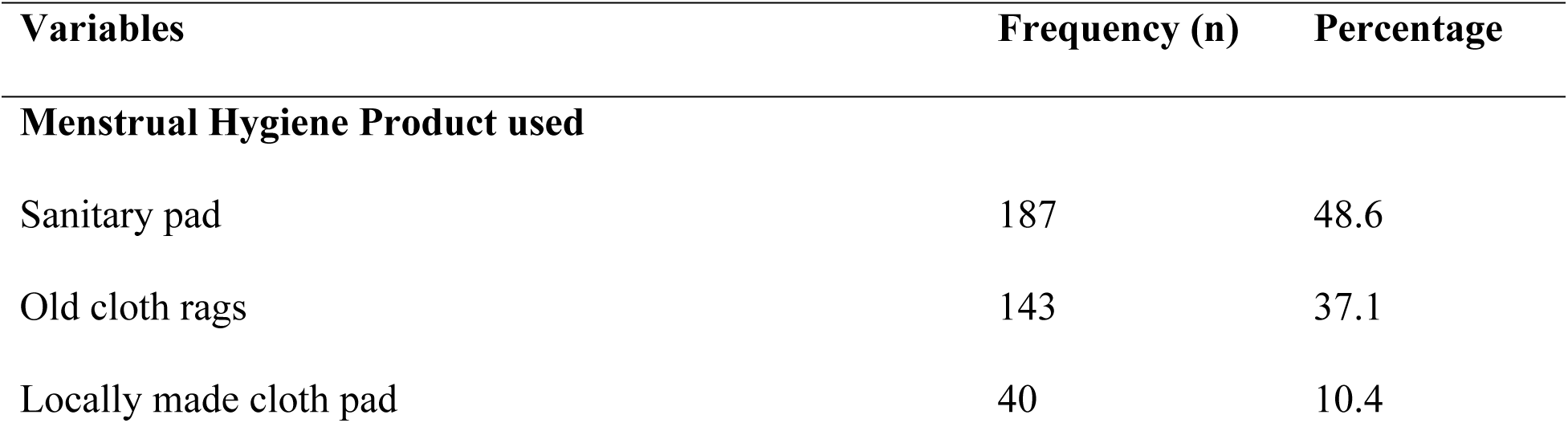
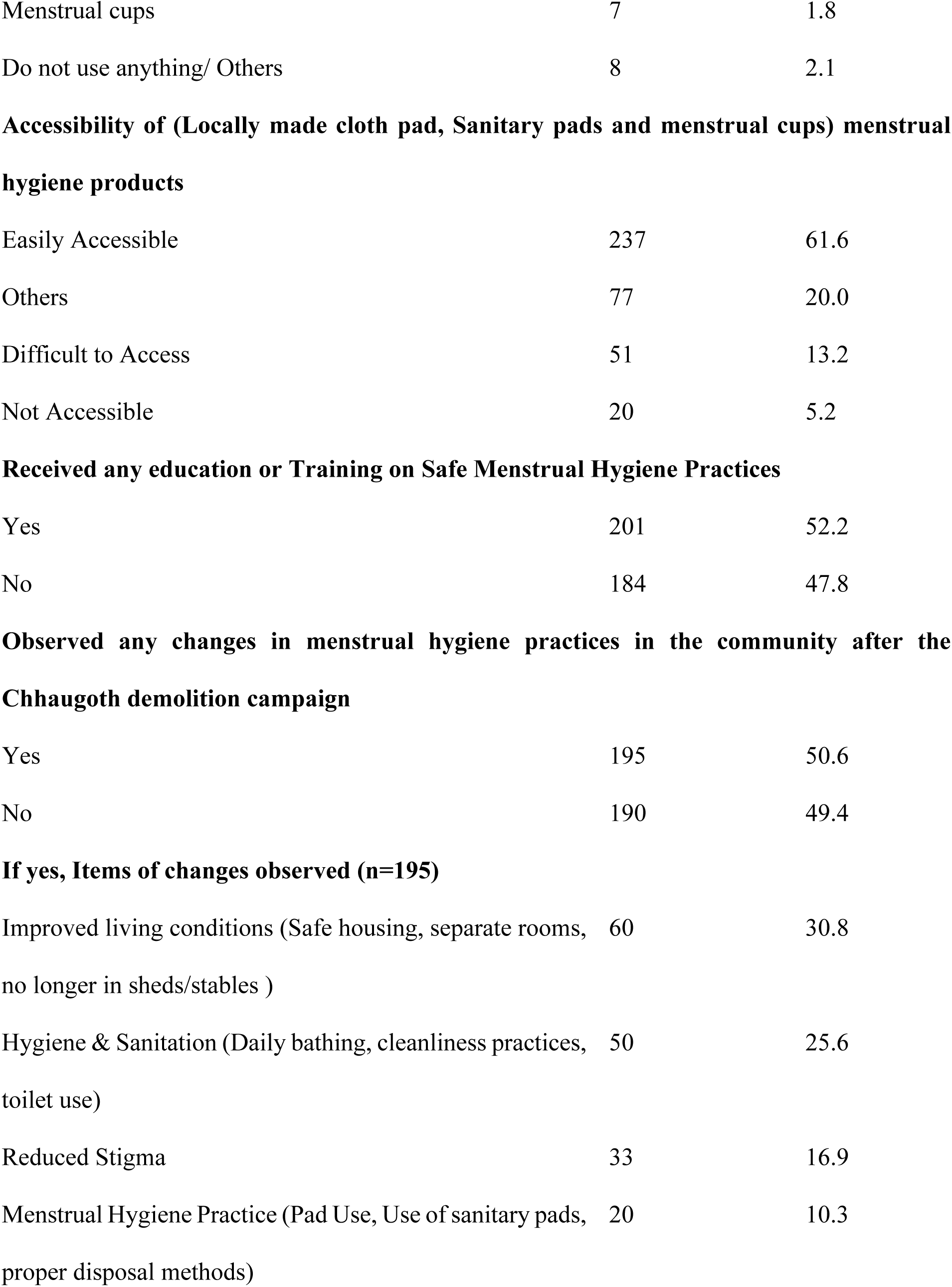
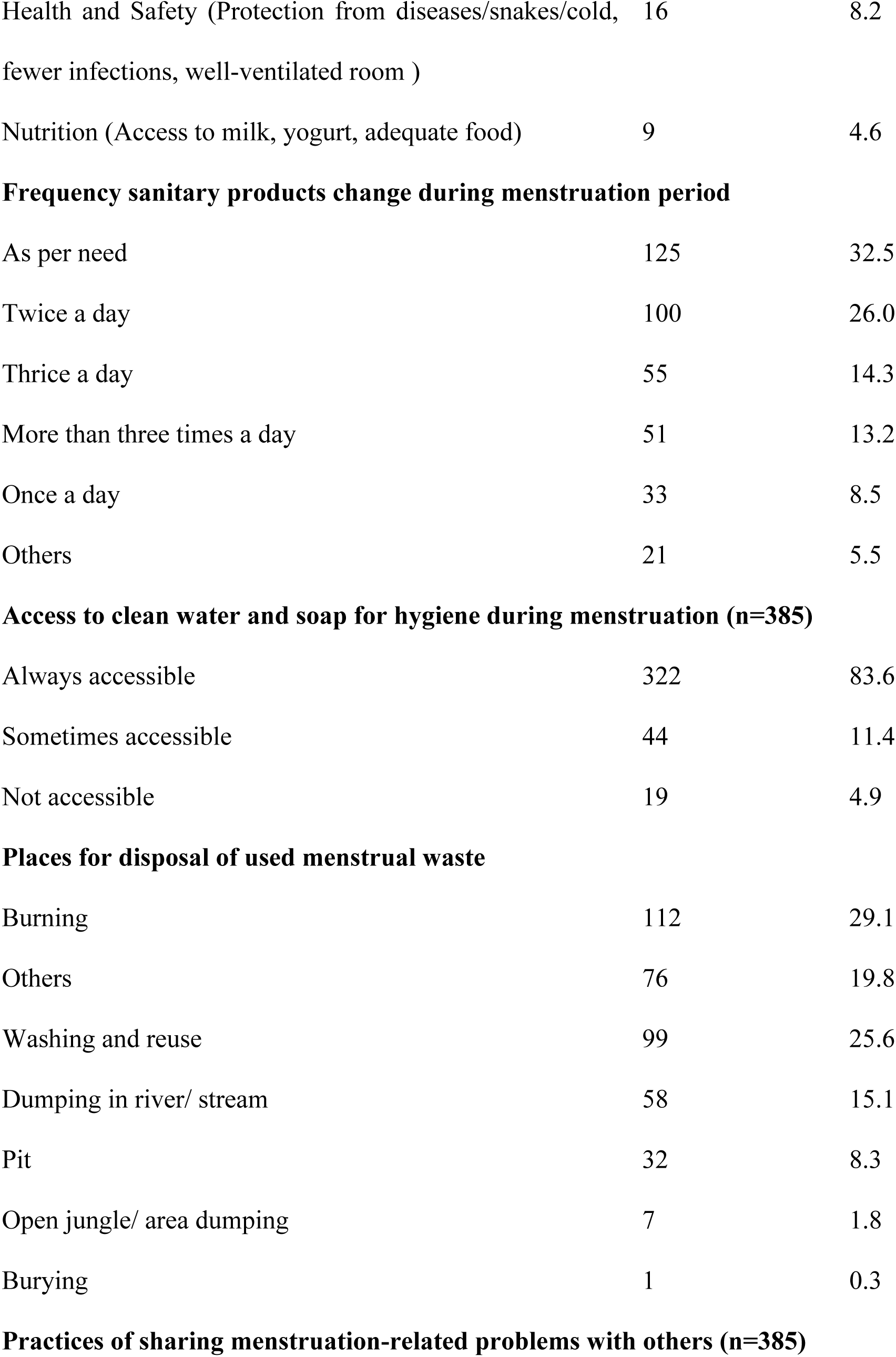
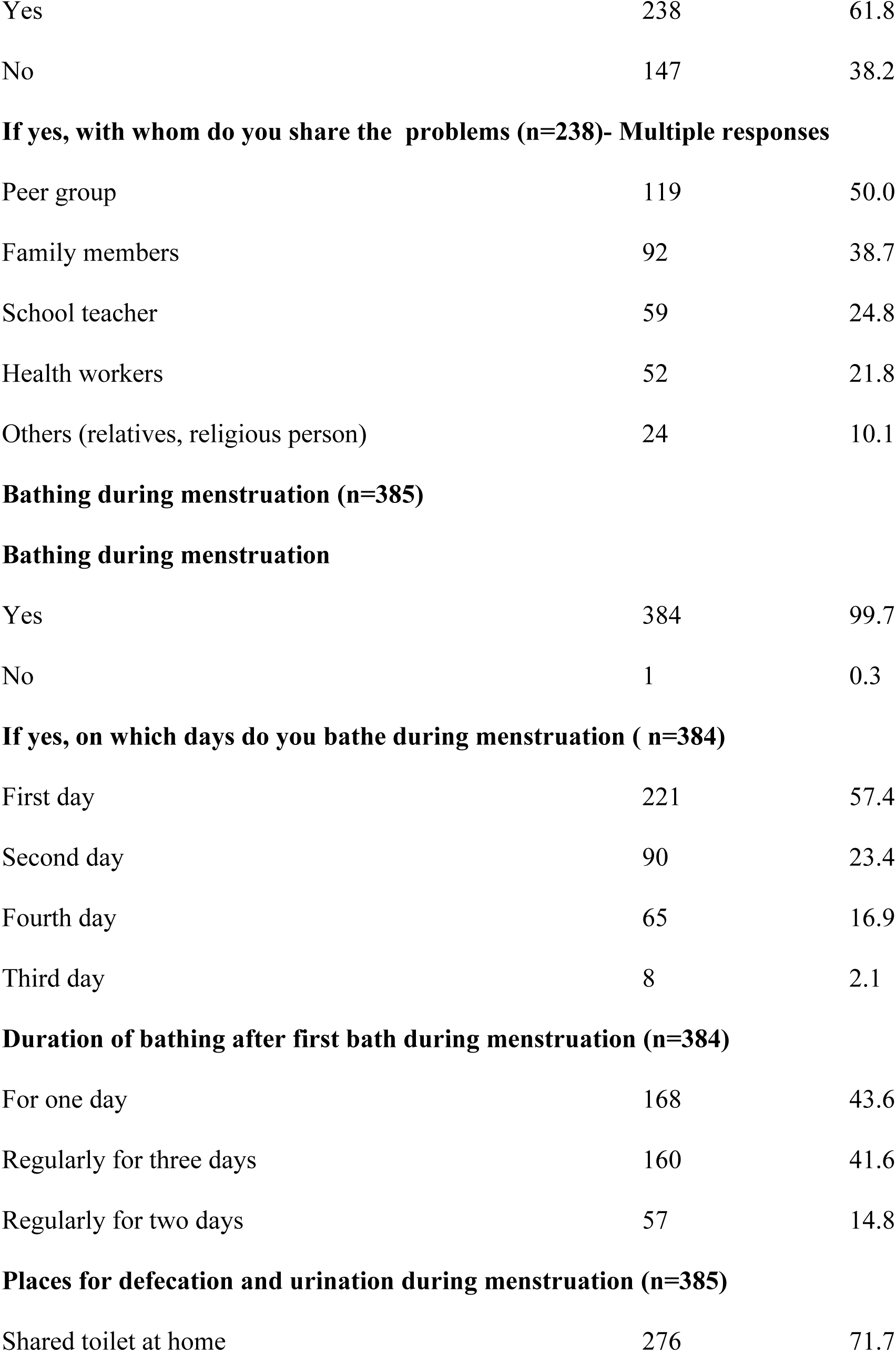
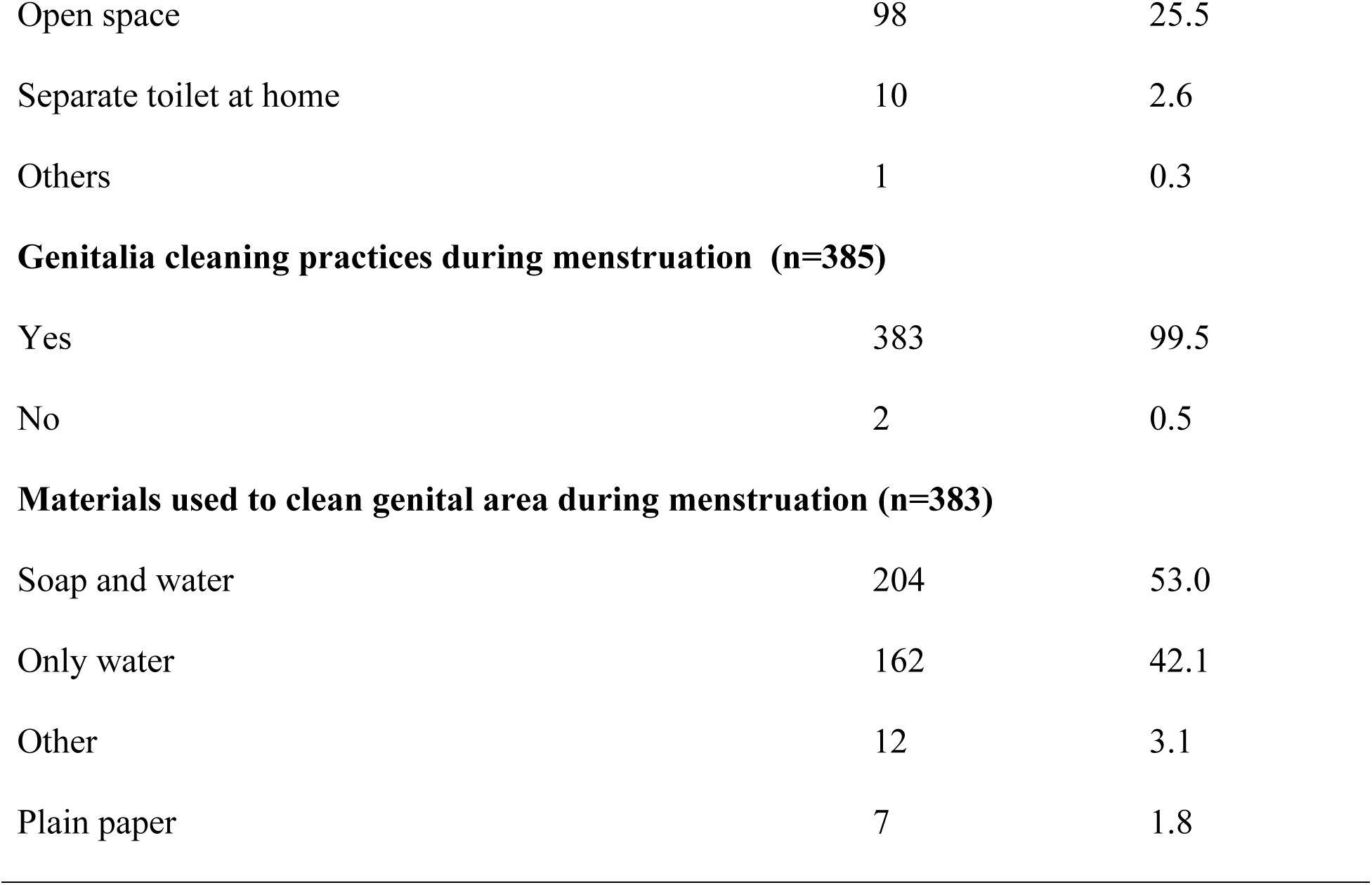
Menstrual Hygiene Management related information (n=385)

| Variables | Frequency (n) | Percentage |
| --- | --- | --- |
| **Menstrual Hygiene Product used** |  |  |
| Sanitary pad | 187 | 48.6 |
| Old cloth rags | 143 | 37.1 |
| Locally made cloth pad | 40 | 10.4 |
| Menstrual cups | 7 | 1.8 |
| Do not use anything/ Others | 8 | 2.1 |
| **Accessibility of (Locally made cloth pad, Sanitary pads and menstrual cups) menstrual hygiene products** |  |  |
| Easily Accessible | 237 | 61.6 |
| Others | 77 | 20.0 |
| Difficult to Access | 51 | 13.2 |
| Not Accessible | 20 | 5.2 |
| **Received any education or Training on Safe Menstrual Hygiene Practices** |  |  |
| Yes | 201 | 52.2 |
| No | 184 | 47.8 |
| **Observed any changes in menstrual hygiene practices in the community after the Chhaugoth demolition campaign** |  |  |
| Yes | 195 | 50.6 |
| No | 190 | 49.4 |
| **If yes, Items of changes observed (n=195)** |  |  |
| Improved living conditions (Safe housing, separate rooms, no longer in sheds/stables ) | 60 | 30.8 |
| Hygiene & Sanitation (Daily bathing, cleanliness practices, toilet use) | 50 | 25.6 |
| Reduced Stigma | 33 | 16.9 |
| Menstrual Hygiene Practice (Pad Use, Use of sanitary pads, proper disposal methods) | 20 | 10.3 |
| Health and Safety (Protection from diseases/snakes/cold, fewer infections, well-ventilated room ) | 16 | 8.2 |
| Nutrition (Access to milk, yogurt, adequate food) | 9 | 4.6 |
| **Frequency sanitary products change during menstruation period** |  |  |
| As per need | 125 | 32.5 |
| Twice a day | 100 | 26.0 |
| Thrice a day | 55 | 14.3 |
| More than three times a day | 51 | 13.2 |
| Once a day | 33 | 8.5 |
| Others | 21 | 5.5 |
| **Access to clean water and soap for hygiene during menstruation (n=385)** |  |  |
| Always accessible | 322 | 83.6 |
| Sometimes accessible | 44 | 11.4 |
| Not accessible | 19 | 4.9 |
| **Places for disposal of used menstrual waste** |  |  |
| Burning | 112 | 29.1 |
| Others | 76 | 19.8 |
| Washing and reuse | 99 | 25.6 |
| Dumping in river/ stream | 58 | 15.1 |
| Pit | 32 | 8.3 |
| Open jungle/ area dumping | 7 | 1.8 |
| Burying | 1 | 0.3 |
| **Practices of sharing menstruation-related problems with others (n=385)** |  |  |
| Yes | 238 | 61.8 |
| No | 147 | 38.2 |
| **If yes, with whom do you share the problems (n=238)- Multiple responses** |  |  |
| Peer group | 119 | 50.0 |
| Family members | 92 | 38.7 |
| School teacher | 59 | 24.8 |
| Health workers | 52 | 21.8 |
| Others (relatives, religious person) | 24 | 10.1 |
| **Bathing during menstruation** |  |  |
| Yes | 384 | 99.7 |
| No | 1 | 0.3 |
| **If yes, on which days do you bathe during menstruation ( n=384)** |  |  |
| First day | 221 | 57.4 |
| Second day | 90 | 23.4 |
| Fourth day | 65 | 16.9 |
| Third day | 8 | 2.1 |
| **Duration of bathing after first bath during menstruation (n=384)** |  |  |
| For one day | 168 | 43.6 |
| Regularly for three days | 160 | 41.6 |
| Regularly for two days | 57 | 14.8 |
| **Places for defecation and urination during menstruation (n=385)** |  |  |
| Shared toilet at home | 276 | 71.7 |
| Open space | 98 | 25.5 |
| Separate toilet at home | 10 | 2.6 |
| Others | 1 | 0.3 |
| **Genitalia cleaning practices during menstruation (n=385)** |  |  |
| Yes | 383 | 99.5 |
| No | 2 | 0.5 |
| **Materials used to clean genital area during menstruation (n=383)** |  |  |
| Soap and water | 204 | 53.0 |
| Only water | 162 | 42.1 |
| Other | 12 | 3.1 |
| Plain paper | 7 | 1.8 |

In most cases (83.6%), the participants reported that their living places during menstruation had windows for ventilation, whereas 16.4% stayed in places without proper ventilation. Majority of the respondents (90.4%) had reported that they had access to locks for privacy; while, more than half of the respondents (56.4%) lacked proper disposal facilities for menstrual waste. Similarly, majority of the respondents (96.1%) had a sleeping place during menstruation. Nearly three quarters of the respondents (73.8%) had regular supply of food while more than a quarter of the respondents (26.2%) faced food restriction during menstruation. Meanwhile, three-quarters of the respondents (75.8%) reported the access to drinking water supply while nearly a quarter of them (24.2%) lacked drinking water access during menstruation. Majority of the respondents (88%) had access to lighting. Meanwhile, more than three quarters of the respondents (79%) had electricity while nearly a quarter of the respondents (21%) lacked electricity access. More than two thirds of the respondents (68.8%) had toilet access while nearly a third (31.2%) lacked access to toilet. Similarly, majority of the respondents (88.3%) had no separate toilet specially for use during their menstrual period. More than half of the respondents (57.9%) stayed within 50 meters, followed 23.7% stayed at a distance of > 100 meters away and 18.4% stayed within 50-100 meters far from the residence; and majority of them (88.1%) reported the facility for space to dry menstrual cloths. (**Table 4**)

**Table 4:** Findings of Observation.

| SN | Observation items | Response | Frequency (n) | Percentage (%) |
| --- | --- | --- | --- | --- |
| 1 | Availability of ventilation/windows in living spaces | Yes | 322 | 83.6 |
| 2 | Availability of locks on doors and windows | Yes | 348 | 90.4 |
| 3 | Presence of place to dispose of used pads | Yes | 168 | 43.6 |
| 4 | Availability of sleeping arrangement in the living space | Yes | 370 | 96.1 |
| 5 | Availability of regular food in living space | Yes | 284 | 73.8 |
| 6 | Availability of drinking water in living space | Yes | 292 | 75.8 |
| 7 | Lighting arrangement inside living space | Yes | 339 | 88 |
| 8 | Availability of electricity in living space | Yes | 304 | 79 |
| 9 | Availability of toilet in living space | Yes | 265 | 68.8 |
| 10 | If yes, is there a separate toilet specifically for use during menstruation (n=304) | Yes | 25 | 9.4 |
|  |  | No | 234 | 88.3 |
|  |  | Yes but others also use | 6 | 2.3 |
| 11 | Distance of the living space from nearby houses | < 50 meters | 223 | 57.9 |
|  |  | 50–100 meters | 71 | 18.4 |
|  |  | > 100 meters | 91 | 23.7 |
| 12 | Availability of space to dry menstrual clothes | Yes | 339 | 88.1 |

## Discussion

This study assessed the living arrangements and menstrual hygiene management (MHM) practices of participants following the government led Chhaugoth demolition campaigns in Chaurpati Rural Municipality, Achham district of Nepal. These findings reveal substantial progress in the living arrangements and hygiene behaviors.

This study reported a remarkable decline in the Chhaugoth use, where a majority of the respondents (93.5%) reported the demolition of their Chhaugoths after the campaign. The decline in Chhaugoth use from 74.8% before to 11.4% after the demolition leading to significant improvement in the living arrangements which suggests behavioral shift and reflect the appreciable outcomes of the demolition campaign, where women and girls are protected and feel safe moving from isolated and hazardous sheds to within their homes. However, 78.2% of the respondents still face cultural taboos and restrictions during their menstrual period and are bounded to follow the seclusion practices within their home. This findings aligns with the studies from the previous studies from different areas of Nepal (1,29,30); where women are secluded in livestock sheds or any specific rooms, which confirms the existence of the practice becoming a silent but impactful cultural restriction. Furthermore, observational data indicates an increasing refinement but not a complete improvement, where still 16.4% of the respondents lacked proper ventilation menstrual waste disposal (56.4%), and consistent supply to safe and clean drinking water (24.2%) and food (26.2%). While compared to dreadful conditions of the traditional Chhaugoths reported by the study of Amatya (1) which indicated a slight improvement but this findings highlights that the current alternate seclusion rooms are not universally safe nor dignified.

Additionally, 48.6% of respondents reported the use of sanitary pads which in comparison to other studies that reported much reliance to cloths/rags as an absorbent materials (1,29). This increasing change could be due to the public awareness about reproductive health problems caused by unsanitary hygiene behaviors related to menstrual hygiene practice. Likewise, majority of the respondents feel comfortable in sharing menstruation related problems with others primarily to the peers (50%) and family members (38.7%). This finding aligns with other studies where their first source of information to menstruation is from their friends and family specifically their mother or sisters (3,20). These findings signify a cultural shift towards openness and normalization of menstruation without feeling shame, where this silence often prevents women from seeking health support during menstruation. Despite such progress, unsafe and unreliable menstrual waste disposal practices remain unchanged. In this study, burning (29.1%) and directly dumping in rivers (15.1%) are the common methods to disposal which is environmentally harmful and can pose multiple health risks. This findings is consistent to other studies of Kanchanpur (31) and Pyuthan (3) districts of Nepal, where the disposal are similar to the current practices of Achhami women and girls. These similarities in findings are mostly due to the fact that these are geographically and culturally connected areas of Sudurpaschim province suggesting regional norms and attributed to these regions shared cultural traditions regarding menstrual impurity and significant migration flows within the province.

This study has some limitations such as its cross sectional nature limits the representation of the relationship between Chhaugoth demolition campaign and the observed changes. Further, the study was conducted in a rural municipality which limits its generalizability and the data collected through interview may lead to response bias as it is of sensitive nature issue. Exclusion of ward no. 7 due to the lack of information related to Chhaugoth demolition campaign could resulted in some sorts of selection bias.

## Conclusion

This study demonstrates remarkable progress in shifting from unsafe, unhygienic, hazardous menstrual seclusion to safer, in-house accommodations; reflecting positive changes following successful Chhaugoth demolition interventions. Living arrangements have improved with more girls and women now residing in secure and separate rooms during menstruation. However, critical gaps remain in menstrual hygiene management (MHM) practices, particularly regarding unsafe menstrual waste disposal and inadequate menstrual friendly infrastructures. This continues to impede safe and dignified MHM. There is the need to strengthen community awareness through targeted Behavior Change Communication (BCC) with the active involvement of family members to eliminate cultural taboos and norms and promote supportive environment. Furthermore, integrating menstrual health education before menarche, equipping girls with early knowledge, awareness and confidence to adopt safe hygiene practices. In addition, these early targeted interventions can foster generational change enabling future generation girls and women to achieve menstrual dignity and equity.

## Data Availability

All relevant data are within the manuscript and its Supporting Information files.

## Acknowledgements

Authors are thankful to the authorities of Chaurpati Rural Municipality, the local Police staff and the study participants in Achham, Nepal, for their invaluable cooperation.

## REFERENCES

1. Amatya P, Ghimire S, Callahan KE, Baral BK, Poudel KC. Practice and lived experience of menstrual exiles (Chhaupadi) among adolescent girls in far-western Nepal. PLoS One. 2018 Dec 10;13(12):e0208260.

2. Shrestha E. Everything you need to know about Chhaupadi, the taboo ritual of banishing women to period huts. The Kathmandu Post [Internet]. 2019 Dec 11; Available from: https://kathmandupost.com/national/2019/12/11/everything-you-need-to-know-about-chhaupadi-the-taboo-ritual-of-banishing-women-to-period-huts

3. Parajuli SB, Kc H, Mishra A, Bhattarai P, Shrestha M, Srivastav K. Chaupadi during menstruation still a major community health challenge: perspective from Mid-Western Nepal. BIBECHANA. 2018 Nov 22;16:228–35.

4. Sommer M, Sahin M. Overcoming the Taboo: Advancing the Global Agenda for Menstrual Hygiene Management for Schoolgirls. Am J Public Health. 2013 Sept;103(9):1556–9.

5. Kadariya S, R. Aro A. Chhaupadi practice in Nepal &ndash; analysis of ethical aspects. MB. 2015 June;53.

6. Karki KB, Poudel PC, Rothchild J, Pope N, Bobin NC, Gurung Y, et al. SCOPING REVIEW AND PRELIMINARY MAPPING Menstrual Health and Hygiene Management in Nepal. 2017 June;

7. Robinson H. *Chaupadi*: The affliction of menses in Nepal. International Journal of Women’s Dermatology. 2015 Dec 1;1(4):193–4.

8. Adhikari A. Stringent measures to end Chhaupadi. The Rising Nepal [Internet]. 2019 Dec 25; Available from: https://old.risingnepaldaily.com/main-news/stringent-measures-to-end-chhaupadi

9. Lamsal A. Engaging Men and Boys Against the Practice of CHHAUPADI in Nepal [Internet]. Centre for Health and Social Justice (CHSJ); 2017 p. 8. Available from: https://www.alignplatform.org/sites/default/files/2020-06/south_asia_case_study_1_engagingmenagainstchhaupadi_practiceinnepal.pdf

10. National Planning Commission Secretariat G of N. Achham Profile Data 2011 [Internet]. 2014 Mar. Available from: https://docs.censusnepal.cbs.gov.np/Documents/eed46218-a3bc-465a-8199-fb325f5e77e369%20Achham_VDCLevelReport.pdf

11. Acharya A, Yadav K, Nongkynrih B. Reproductive Tract Infections/ Sexually Transmitted Infections in Rural Haryana: Experiences from the Family Health Awareness Campaign. Indian Journal of Community Medicine. 2006 Jan 1;31.

12. Ranabhat C, Kim CB, Choi EH, Aryal A, Park MB, Doh YA. Chhaupadi Culture and Reproductive Health of Women in Nepal. Asia Pac J Public Health. 2015 Oct 1;27(7):785–95.

13. Bhatta B. Woman dies of snakebite in menstrual shed in Kanchanpur. The Kathmandu Post [Internet]. 2025 Sept 15; Available from: https://kathmandupost.com/sudurpaschim-province/2025/07/13/woman-dies-of-snakebite-in-menstrual-shed-in-kanchanpur

14. the Himalayan Times the HT. Chhaupadi claims two more lives in Achham. The Himalayan Times. 2025 Aug 29;

15. Kaur R, Kaur K, Kaur R. Menstrual Hygiene, Management, and Waste Disposal: Practices and Challenges Faced by Girls/Women of Developing Countries. Journal of Environmental and Public Health. 2018;2018(1):1730964.

16. Sumpter C, Torondel B. A Systematic Review of the Health and Social Effects of Menstrual Hygiene Management. RezaBaradaran H, editor. PLoS ONE. 2013 Apr 26;8(4):e62004.

17. Dailymail.com. Mail Online. 2018 [cited 2024 Sept 4]. Nearly half of American women have been “period SHAMED.” Available from: http://www.dailymail.co.uk/femail/article-5233629/Nearly-half-American-women-period-SHAMED.html

18. Martinet A. Menstruation, taboo within the couple for three-quarters of French women. Le Journaldes Femmes Love [Internet]. 2017; Available from: https://www.journaldesfemmes.fr/couple/vie-a-deux/1814781-regles-sujet-tabou/

19. The Period Taboo: A Universal Problem | CARE International [Internet]. 2021 [cited 2024 Sept 4]. Available from: https://www.care-international.org/stories/period-taboo-universal-problem

20. Mukherjee A, Lama M, Khakurel U, Jha AN, Ajose F, Acharya S, et al. Perception and practices of menstruation restrictions among urban adolescent girls and women in Nepal: a cross-sectional survey. Reproductive Health. 2020 June 1;17(1):81.

21. Nguyen V. UNDP bangladesh. 2022 [cited 2024 Sept 4]. Meet the Women Fighting to End Period Stigma in Rural Bangladesh. Available from: https://www.undp.org/bangladesh/blog/meet-women-fighting-end-period-stigma-rural-bangladesh

22. Khanna A, R.S. Goyal, Bhawsar R. Menstrual Practices and Reproductive Problems: A Study of Adolescent Girls in Rajasthan. Journal of Health Management. 2005 Apr;7(1):91–107.

23. UNFPA UNPF. Menstruation and human rights - Frequently asked questions | United Nations Population Fund [Internet]. 2022 [cited 2024 Sept 20]. Available from: https://www.unfpa.org/menstruationfaq

24. WHO WHO. Menstrual health is a fundamental human right [Internet]. 2024 [cited 2024 Sept 20]. Available from: https://www.who.int/europe/news/item/15-08-2024-menstrual-health-is-a-fundamental-human-right

25. Sommer M, Caruso BA, Sahin M, Calderon T, Cavill S, Mahon T, et al. A Time for Global Action: Addressing Girls’ Menstrual Hygiene Management Needs in Schools. PLOS Medicine. 2016 Feb 23;13(2):e1001962.

26. Dhungana M. Chhaugoth was destroyed, Chhaupadi was not destroyed. The Kantipur post [Internet]. 2024 May 8; Available from: https://ekantipur.com/en/news/2024/04/20/chhaugoth-was-destroyed-chhaupadi-was-not-destroyed-17-47.html#:~:text=At%20that%20time,%20according%20to%20the%20Ministry’s%20circular,%20District%20Police

27. Ghimire A. Menstrual Exile: Nepal’s Chhaupadi and the Policy-Practice Divide. Int J Public Health. 2024 Dec 16;69:1608202.

28. Rising Nepal. Fight Against Chhaupadi. The Rising Nepal [Internet]. 2019 Dec 26; Available from: https://old.risingnepaldaily.com/editorial/fight-against-chhaupadi#

29. Thakuri DS, Thapa RK, Singh S, Khanal GN, Khatri RB. A harmful religio-cultural practice (Chhaupadi) during menstruation among adolescent girls in Nepal: Prevalence and policies for eradication. PLoS One. 2021 Sept 1;16(9):e0256968.

30. Thapa S, Bhattarai S, Aro AR. “Menstrual blood is bad and should be cleaned”: A qualitative case study on traditional menstrual practices and contextual factors in the rural communities of far-western Nepal. SAGE Open Med. 2019;7:2050312119850400.

31. Lama S. Experiences of menstrual exiles (Chaupadi) and its consequences among women in Nepal. 2019 [cited 2025 July 2]; Available from: https://digital.car.chula.ac.th/cgi/viewcontent.cgi?article=9844&context=chulaetd

